# Public Attitudes Towards Alcohol and Alcohol Taxation Policies: Insights from a Cross-National Survey

**DOI:** 10.1101/2025.09.18.25333312

**Authors:** Nandita Murukutla, Nalin Singh Negi, Rachel Rothenstein-Henry, Meena Maharjan, Andrew Johnson, Jacqui Drope, Sandra Mullin, Adam Karpati

## Abstract

**Aims:** Alcohol consumption is a major global health risk, yet effective policies like alcohol taxation remain politically contentious and underutilized. Since public support is critical for advancing these measures, this study explores public attitudes towards alcohol harms and taxation in Brazil, Colombia, Mexico, Kenya, and the Philippines, revealing opportunities and barriers for immediate policy action.

**Design, Setting, and Participants:** Nationally representative cross-sectional surveys among adults aged 18+ years, conducted between March 8 and April 15, 2024, using telephone surveys in Brazil, Colombia, and Mexico and in-person household surveys in Kenya and the Philippines (Ns=1000–1096 per country).

**Measurements:** Key outcome measures were attitudes towards alcohol tax policies. Predictors included country of residence, demographic characteristics, alcohol use status, exposure to media coverage of alcohol harms, concern about and knowledge of alcohol-related harms, attitudes toward government responsibility in addressing alcohol harms, perceptions of alcohol industry interference in policymaking, and industry culpability for alcohol-related damage.

**Findings:** Substantial majorities (88% overall; range: 66% Philippines–94% Colombia, and Mexico) identified alcohol consumption as a problem, with violence as the most prominent concern (55% overall; range: 38% Mexico–62% Colombia). Majorities agreed that it is the government’s responsibility to address alcohol’s harms (79% overall; range: 66% Philippines–89% Kenya); that the alcohol industry is culpable for these harms (62% overall; range: 59% Brazil and Philippines–71% Kenya); and that alcohol taxes are an effective policy measure (65% overall; range: 55% Mexico–67% Philippines). In regression analyses, predictors of agreement in alcohol tax effectiveness included: media exposure to alcohol harms (AOR=1.18, 95% CI:1.03–1.36); alcohol use status (AOR=0.74, 95% CI:0.62–0.89); recognition of the population-wide benefits of alcohol policies (AOR=1.56, 95% CI:1.36–1.79); beliefs in government responsibility (AOR=1.70, 95% CI:1.47–1.97), alcohol industry interference in policymaking (AOR=1.43, 95% CI:1.25–1.64), and alcohol industry culpability for for alcohol-related damages (AOR=1.46, 95% CI:1.28–1.66).

**Conclusions:** Public concern about alcohol harms, especially violence, is widespread, underscoring the urgent need for policy action. While alcohol taxation is recognized as an effective policy solution by majorities in all countries, policymakers must clearly communicate their broader societal benefits, reinforce the government’s responsibility in protecting public health, and explicitly expose alcohol industry culpability and interference in policymaking to strengthen and sustain public support.

## INTRODUCTION

Alcohol consumption is a significant global health threat, contributing to 2.6 million deaths annually (1,2), increasing the risks for non-communicable diseases like liver disease, heart disease, and cancer, and leading to violence, road crashes, falls, and injuries. Its economic costs are considerable, with direct healthcare expenses and productivity losses estimated at 2 to 3% of a country’s gross domestic product (GDP) (3). Effective alcohol policy solutions are available in the World Health Organization’s SAFER technical package, including alcohol taxation to raise prices and reduce consumption. Yet, while global adoption of these measures improved between 2010 and 2019, the most effective measures remain underutilized. For example, although countries impose alcohol excise taxes, these taxes often exclude (or are applied at lower rates) to many products, fail to adjust for inflation, and are set too low to meaningfully improve public health (4,5)

A major obstacle to effective alcohol policies is the alcohol industry’s multi-faceted strategy to glamorize drinking, minimize perceived harms, and obstruct evidence-based policy, mirroring historical tobacco industry tactics (6–8). Alcohol companies target youth and other vulnerable populations through sophisticated marketing that normalizes consumption and downplays chronic alcohol-related diseases, while employing pseudo-scientific research, “corporate social responsibility” campaigns, and media narratives to promote self-regulation, deflect scrutiny, and maintain lenient regulatory environments (9–12). Exposing these manipulative practices and highlighting industry interference is crucial for building public support for comprehensive alcohol control policies.

### Public opinion and policy success

Public support is crucial in advancing and safeguarding effective alcohol policies (13). However, several studies have shown that broader, more impactful measures—such as alcohol taxation or restrictions on availability—receive lower public support than narrower, less-effective interventions targeting specific subpopulations, such as treatment programs for alcohol use disorders (14–16). This pattern likely stems from industry-driven narratives that exploit public anxieties about taxes and rising prices, manipulating these public concerns into resistance specifically against alcohol taxes (6,9,17). Such narratives undermine political willingness to implement these proven, cost-effective policies.

Despite these challenges, public pressure remains a powerful driver for government action, compelling policymakers to introduce new policies or strengthen enforcement of existing regulations (18–20). Evidence indicates that support for policies, especially complex fiscal measures like excise taxes, increases when the public understands their effectiveness (21,22). Consequently, shaping public perception of alcohol as a pressing health issue and improving awareness of the benefits of “best buy” policies are critical for success (23). When policymakers recognize broad public backing, it can bolster their pursuit of effective policies that might otherwise seem too controversial or politically risky (18–20).

Additionally, public awareness of industry interference significantly strengthens public demand for policy action. Evidence shows that when people are informed about manipulative industry tactics designed to delay or weaken policies, their opposition to the industry grows, and support for public health policies increases (11,24,25). These findings underscore the importance of industry denormalization efforts—i.e., exposing industry tactics and harms to change public perception and emphasize that tobacco use is not a mainstream activity— in building consensus for taxation and other policy measures to reduce alcohol-related harm (14,26–30).

Launched in 2022, RESET Alcohol aims to implement WHO’s SAFER package policies, prioritizing alcohol taxation. Recognizing the importance of public attitudes, multi-country surveys were conducted in Brazil, Colombia, Mexico, Kenya and the Philippines to assess public awareness and prioritization of alcohol-related harms, perceptions of alcohol industry influence, support for government action on alcohol policies, and attitudes towards alcohol taxation, including associated socio-demographic and attitudinal predictors.

## METHODS

Nationally representative cross-sectional surveys were conducted by Thinks Insight, a research consultancy, and a network of local field partners in Brazil, Colombia, Mexico, Kenya and the Philippines between March 8, 2024, and April 15, 2024.

### Study participants

Each survey sample comprised adults aged 18 and older (Ns: Brazil = 1001; Colombia = 1003; Mexico = 1006; Kenya = 1096; and the Philippines = 1000). Sampling quotas for socio-demographic criteria were established in collaboration with local survey teams to ensure a high-quality, representative sample, based on available statistics in each country and the data collection methods used. The criteria included region (in Brazil, Colombia, Mexico, Kenya and the Philippines), gender (in Brazil, Colombia, Mexico and the Philippines), age (in Brazil, Colombia and Mexico), education (in Brazil and Colombia), place of residence (rural/urban) (in Kenya and the Philippines), and parents with children (in all five countries). Quotas for parents with children were based on the United Nations Household Size and Composition Estimates from 2022. Individuals working in the tobacco, alcohol or market research industries were excluded.

### Data collection

Cross-sectional surveys were designed to provide nationally representative samples, with sample sizes calculated to yield a margin of error of ±3 percentage points per country. Random-digit-dial telephone surveys were used in Brazil, Colombia and Mexico, with quotas based on regional stratification and proportional sampling from mobile and landline databases. In Kenya and the Philippines, multi-stage face-to-face household surveys employed regional and urban-rural stratification, with households selected by random-route methods and one adult per household randomly chosen using computer-assisted personal interviewing. Up to two revisits were attempted before substitution.

### Questionnaire

The questionnaire, guided by prior literature, assessed public concerns, knowledge, attitudes, and attitudes towards alcohol policies using a standardized instrument and protocol across countries, with minor local adaptation.

Concern about alcohol use was measured by asking, *“In your opinion, how much of a problem is alcohol consumption in [country]?”* (1=not at all, 5=major). Participants also spontaneously listed, without limitation, the most concerning alcohol-related problems: *“Which are the problems related to alcohol consumption in your country that concern you most? What else? Anything else?”* Knowledge of alcohol-related harms was assessed using: *“To the best of your knowledge, does the consumption of alcohol increase the risk of…”* followed by a series of listed diseases, with participants indicating yes/no. The importance of alcohol education was assessed based on agreement with the statement, *“It is important to educate children and adolescents about the risks and consequences of alcohol consumption.”*

The following attitudes were measured using 5-point agreement scales (1=strongly disagree, 5=strongly agree): alcohol accessibility with *“Alcohol is easy to buy”* and *“Alcohol is relatively inexpensive in [country];”* attitudes towards government responsibility were assessed through (dis)agreement with: *“It is the government’s responsibility to address the problems with alcohol consumption in my country”*; *“Our current alcohol laws protect people from alcohol harms”*; *“Our current alcohol laws are poorly enforced/implemented”*; and *“Policy measures to reduce the consumption of alcohol can benefit the public whether they consume alcohol or not.”* Attitudes towards the alcohol industry were captured through (dis)agreement with: *“Alcohol companies can be trusted to tell the truth about the harms of alcohol use”*; and *“The alcohol industry interferes with the passage of new alcohol policies in my country.”* Additionally, attitudes toward industry culpability were assessed using the statement: *“Alcohol companies should take responsibility for the harm caused by alcohol use.”* For clarity, we present this original survey statement in all data tables. However, within the accompanying narrative text, we refer to these attitudes using the term *culpability*, as this provides a more analytically precise framing for interpreting public perceptions of industry accountability.

Attitudes toward alcohol taxation policies were assessed by (dis)agreement with: *“Taxes/increases in taxes on alcohol products would be effective in reducing alcohol consumption”* and *“Increasing the price of alcohol would help reduce alcohol consumption”*

Socio-demographic measures included gender, education, age and urbanity. Alcohol use was measured using WHO STEPS questions: *“Have you ever consumed any alcohol such as beer, wine, spirits or [local examples]?”*; *“Have you consumed any alcohol within the past 12 months?”* and *“Have you consumed any alcohol within the past 30 days?”* Exposure to alcohol harm-related media was assessed via: *“During the last two months, have you seen, read or heard any advertisements, public service announcements or messages in the media about the harms related to alcohol consumption?”*

The questionnaire was translated and back-translated into local languages: Kiswahili (Kenya); Tagalog, Cebuano, Hiligaynon and Waray (the Philippines); Spanish (Colombia and Mexico); and Portuguese (Brazil). The questionnaire was rigorously pilot tested. In this paper, we report only questions directly pertinent to this analysis.

### Data analysis

Nationally representative estimates were generated using random iterative method (RIM) weighting (raking), based on region (Brazil, Colombia, Kenya, Philippines), gender and age (Brazil, Mexico, Kenya), education (Brazil), urban-rural residence (Kenya, Philippines), and socio-economic status (Philippines). Column proportion tests (SPSS v25) compared categorical variables across countries. Ordinal regression (R) identified predictors of favorable attitudes towards alcohol tax policies, using unadjusted and adjusted odds ratios for country, demographics, alcohol use, media exposure, perceptions, knowledge and attitudes. Due to multicollinearity, parental status was excluded, and missing data were listwise deleted. A composite score for perceived alcohol tax effectiveness combined two relevant items (Cronbach’s α=0.754), categorized as low (≤2), medium (>2 to <4), or high (≥4). Other attitudinal items (government responsibility, alcohol industry views) did not show sufficient internal consistency, thus were analysed separately. A knowledge scale summed 15 alcohol-harm awareness items (range: 0–15), categorized as low (0–5), medium (6–10), or high (11–15). The analyses were not pre-registered and the results should be considered exploratory.

## RESULTS

### Sample demographics

Sample demographics (Table 1) showed balanced gender distribution across countries, with Kenya’s participants being youngest and Colombia’s and Mexico’s oldest. Kenya and the Philippines had the highest proportions of participants with children under 18. Exposure to anti-alcohol messaging was lowest in Brazil and Mexico, highest in Kenya. Current alcohol use was highest in Brazil and lowest in Kenya, with survey-based estimates generally lower than WHO prevalence data, especially in Colombia and Kenya.

**Table 1.**
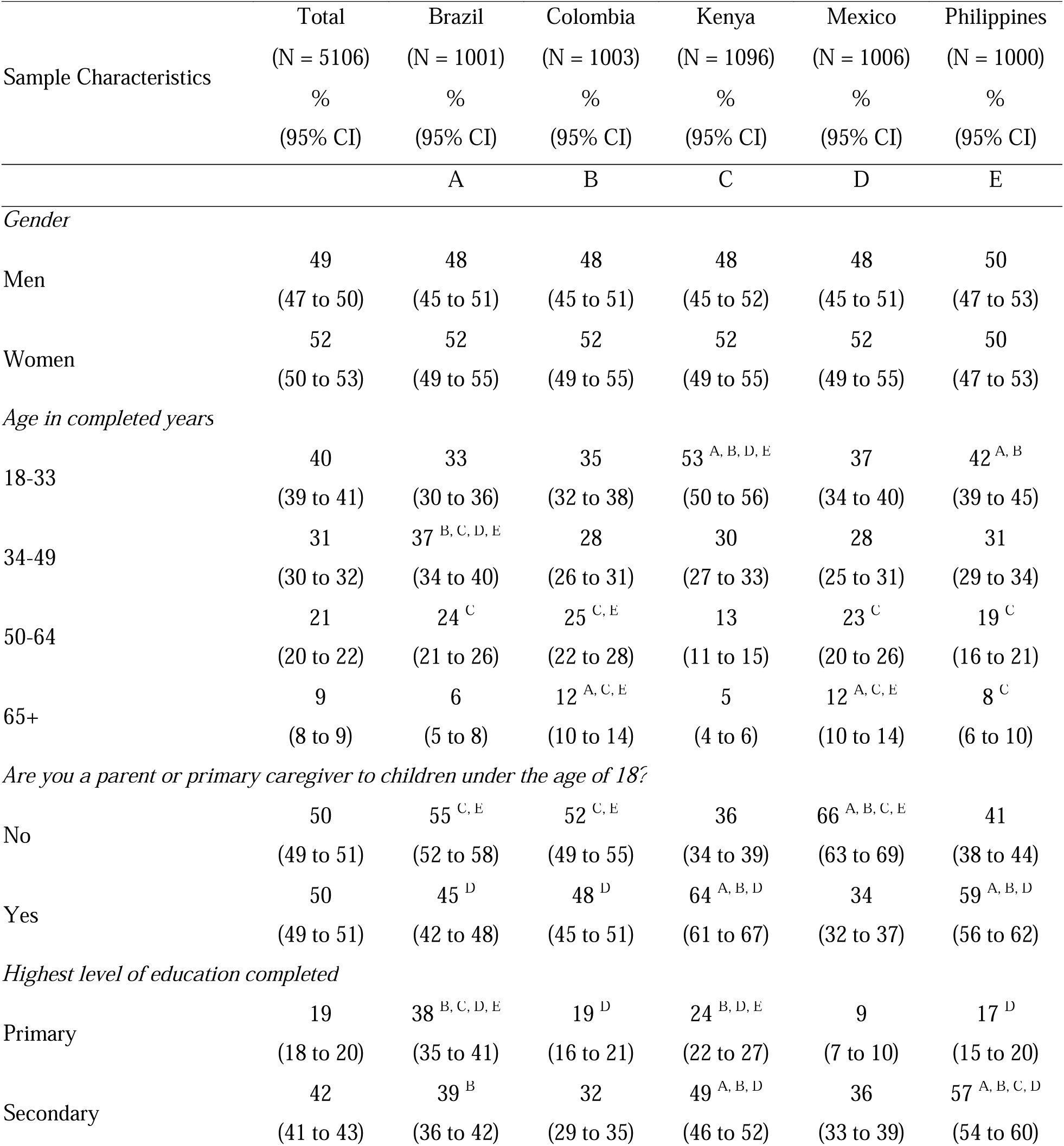
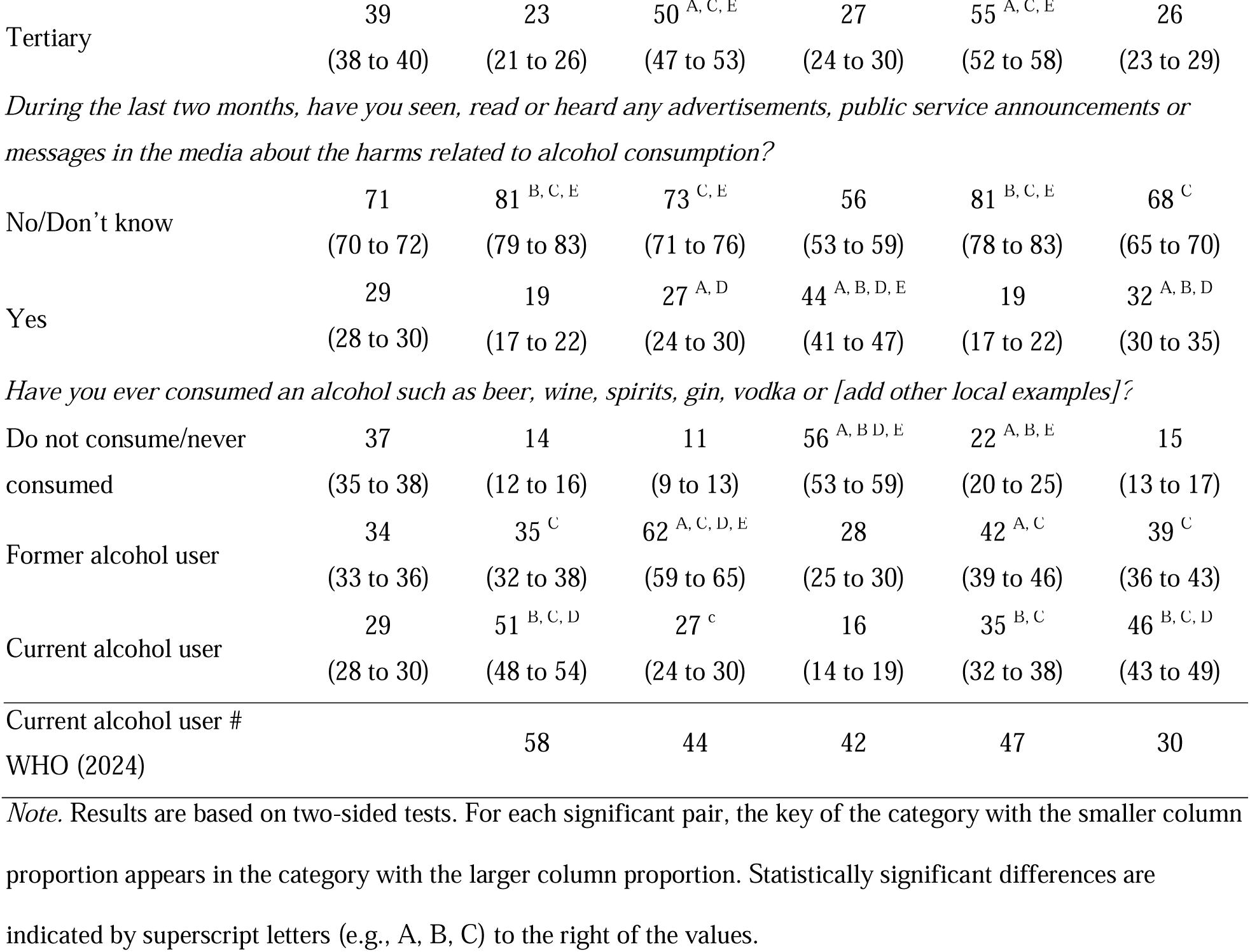
Sample characteristics and alcohol consumption by country.

### Concern about alcohol and knowledge of alcohol harms

Majorities in all countries viewed alcohol consumption as a moderate-to-major problem, highest in Colombia and Mexico and lowest in the Philippines (see Table 2). Among those who reported alcohol as a major to moderate problem, specific top-of-mind concerns varied: violence ranked highest in Colombia, Kenya and the Philippines; health-related problems dominated in Kenya and Brazil, with family and community issues also prominent in Kenya.

**Table 2.**
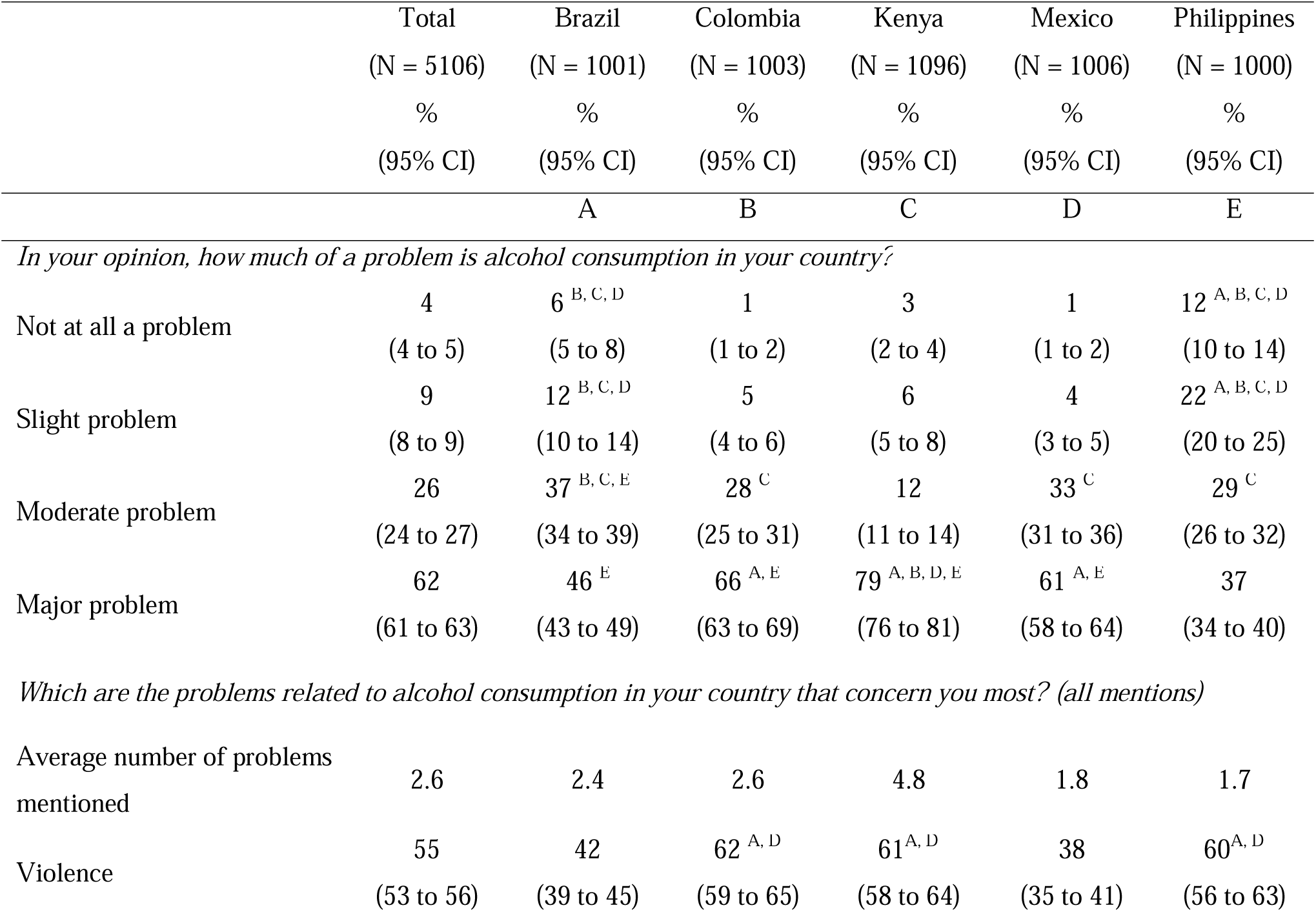
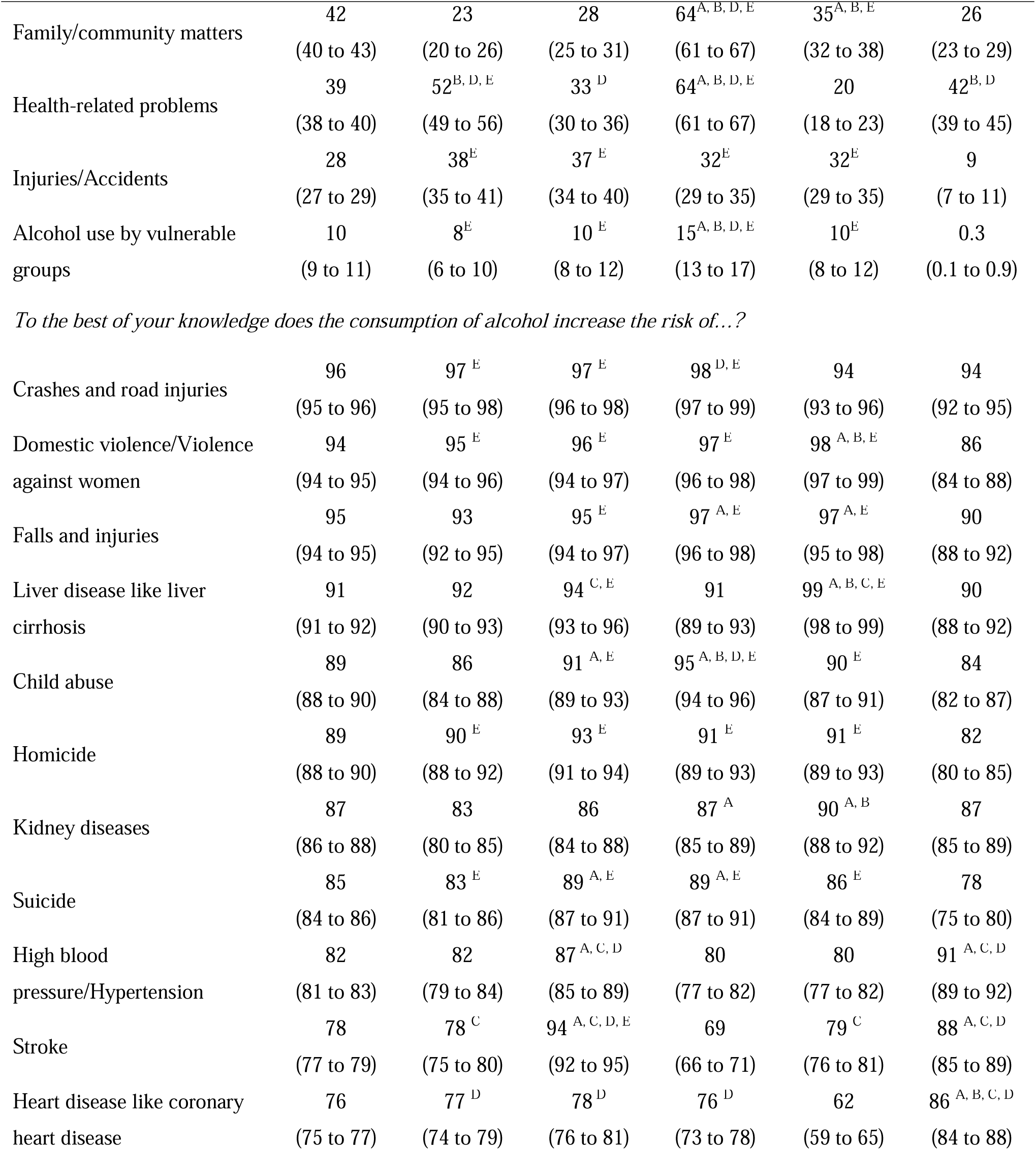
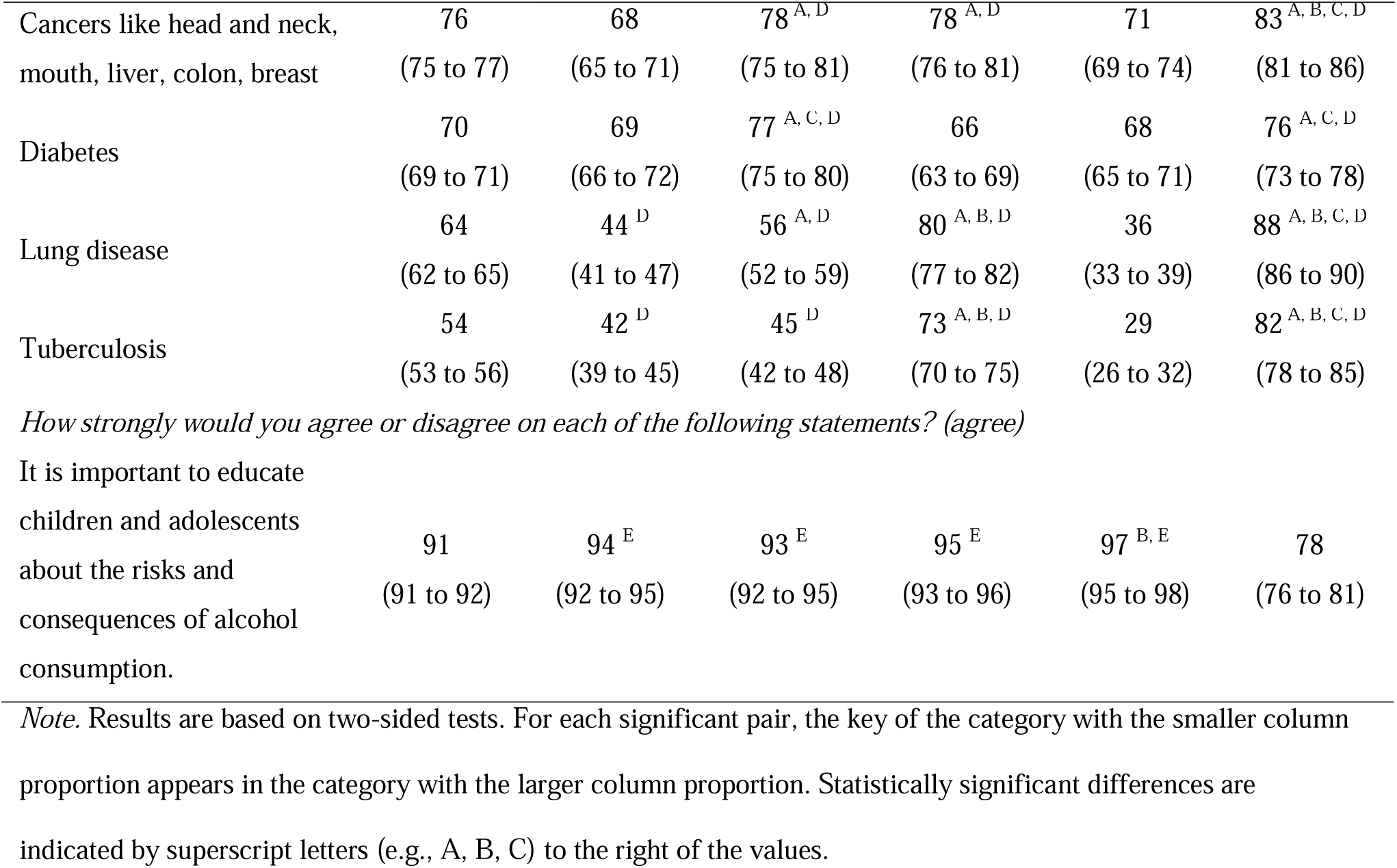
Concerns about and knowledge of alcohol harms by country.

Among health-related concerns mentioned spontaneously, NCDs were cited most frequently in Kenya (35%) and least in Mexico (2%; these percentages are reported here for the context but are not presented in Table 2.). Liver disease, like cirrhosis, was most common concern (Kenya 20%, Brazil 10%), followed by kidney disease (Kenya 11%, Colombia 7%). Other NCDs (cancer, heart disease, stroke, lung disease) were infrequently mentioned (<9% in all countries) (these percentages are reported here for the context but are not presented in Table 2).

Overall knowledge of alcohol-related harms was high (Table 2): highest in Mexico and Kenya, lowest in Brazil and the Philippines, and generally stronger for violence-related harms than NCDs. All countries strongly supported educating youth about alcohol risks: >93% in Mexico, Kenya, Brazil and Colombia and slightly lower in the Philippines (78%).

### Attitudes towards alcohol accessibility, government’s role, and the alcohol industry

Table 3 describes public perceptions of alcohol accessibility, attitudes toward the government, current alcohol policies, and the role of the alcohol industry. Majorities in all countries found alcohol easy to buy and relatively inexpensive, with highest agreement in Brazil and lowest in the Philippines and Kenya. Majorities agreed that it is the government’s responsibility to address alcohol problems, and that policy measures would benefit the public. Only a minority in Brazil, Colombia and Mexico agreed that current laws protect people from alcohol harm; in Kenya and the Philippines, it was about half (49% and 50%, respectively). Majorities across countries agreed that current laws are poorly enforced.

**Table 3.**
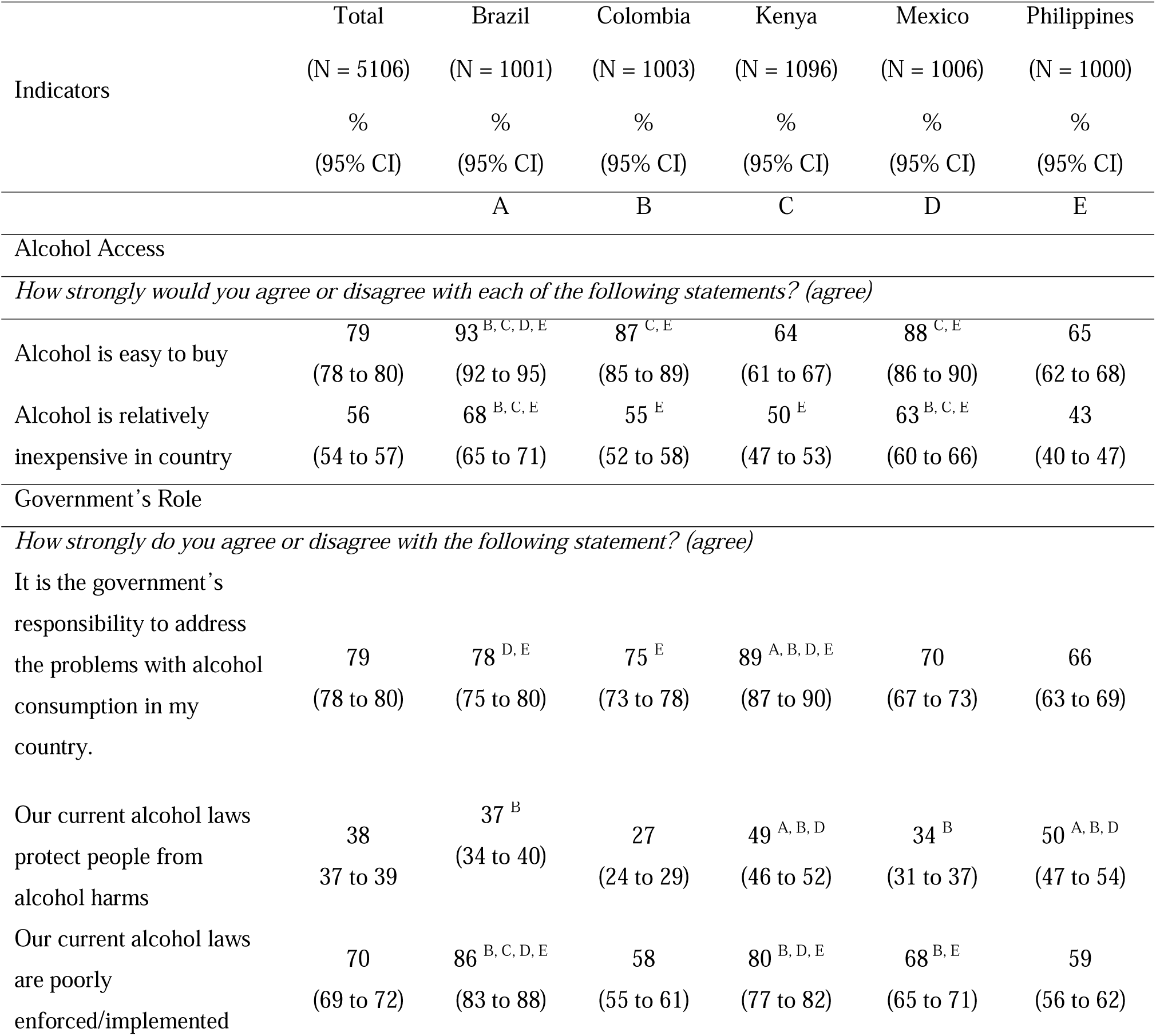
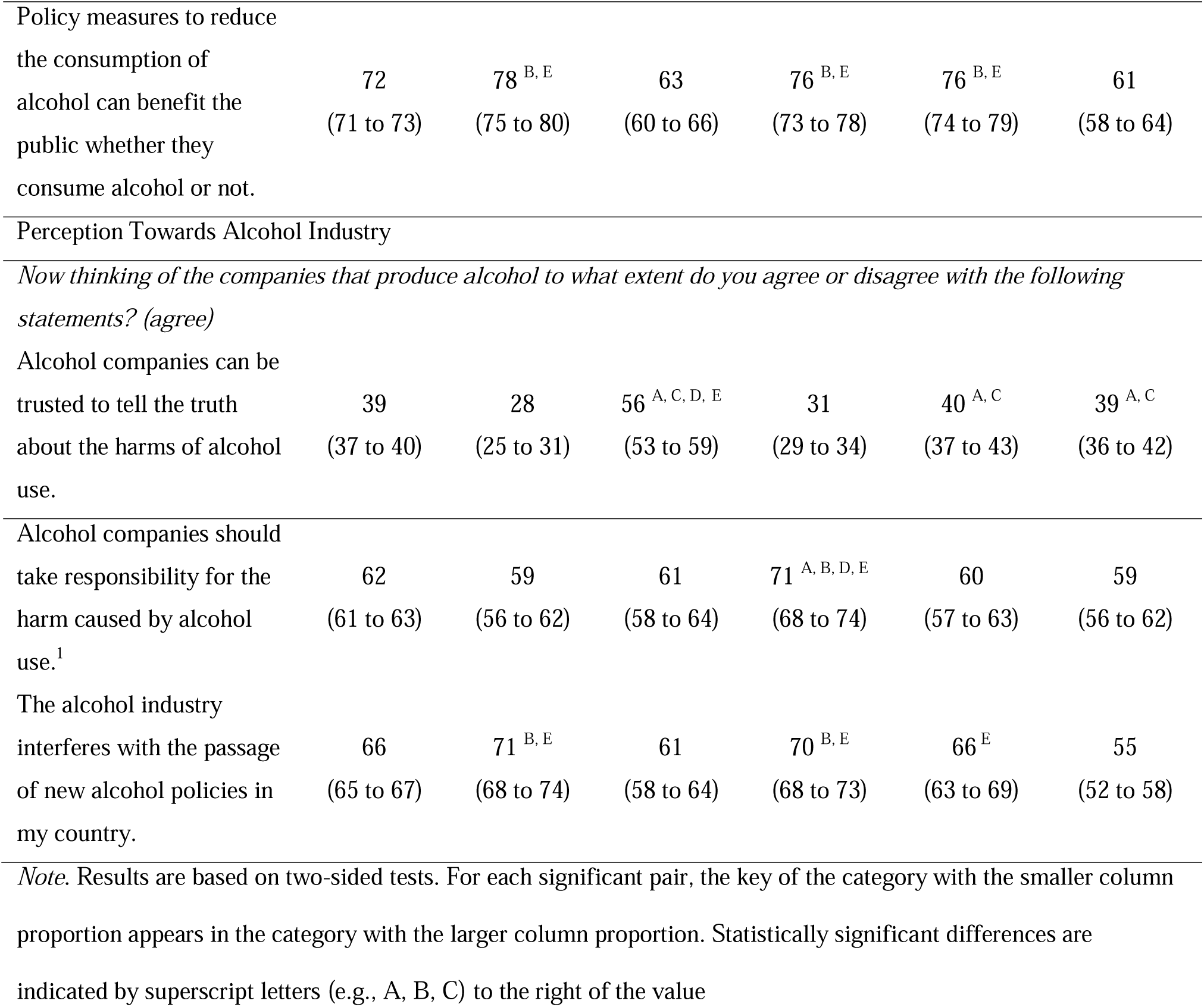
Attitudes towards alcohol accessibility, government’s roles, and the alcohol industry.

Perceptions of the alcohol industry were predominantly negative, except in Colombia (see Table 3). Trust in alcohol companies was low in all countries except Colombia, where a small majority expressed trust. Agreement that alcohol companies are culpable—i.e., that they should take responsibility for alcohol harms—was highest in Kenya, with moderate levels elsewhere, while perceptions of industry interference in policy were highest in Kenya and Brazil, and lowest in Colombia and the Philippines.

### Attitudes towards alcohol taxation policies

Attitudes toward alcohol taxation policies are presented in Table 4. Majorities in all countries agree that taxes on alcohol products would effectively reduce consumption. Likewise, majorities in all countries, except Mexico (where it was 49%) agreed that a higher price on alcohol products would reduce consumption. The most favourable attitudes towards alcohol taxation policies were expressed in the Philippines and the least in Mexico.

**Table 4.**
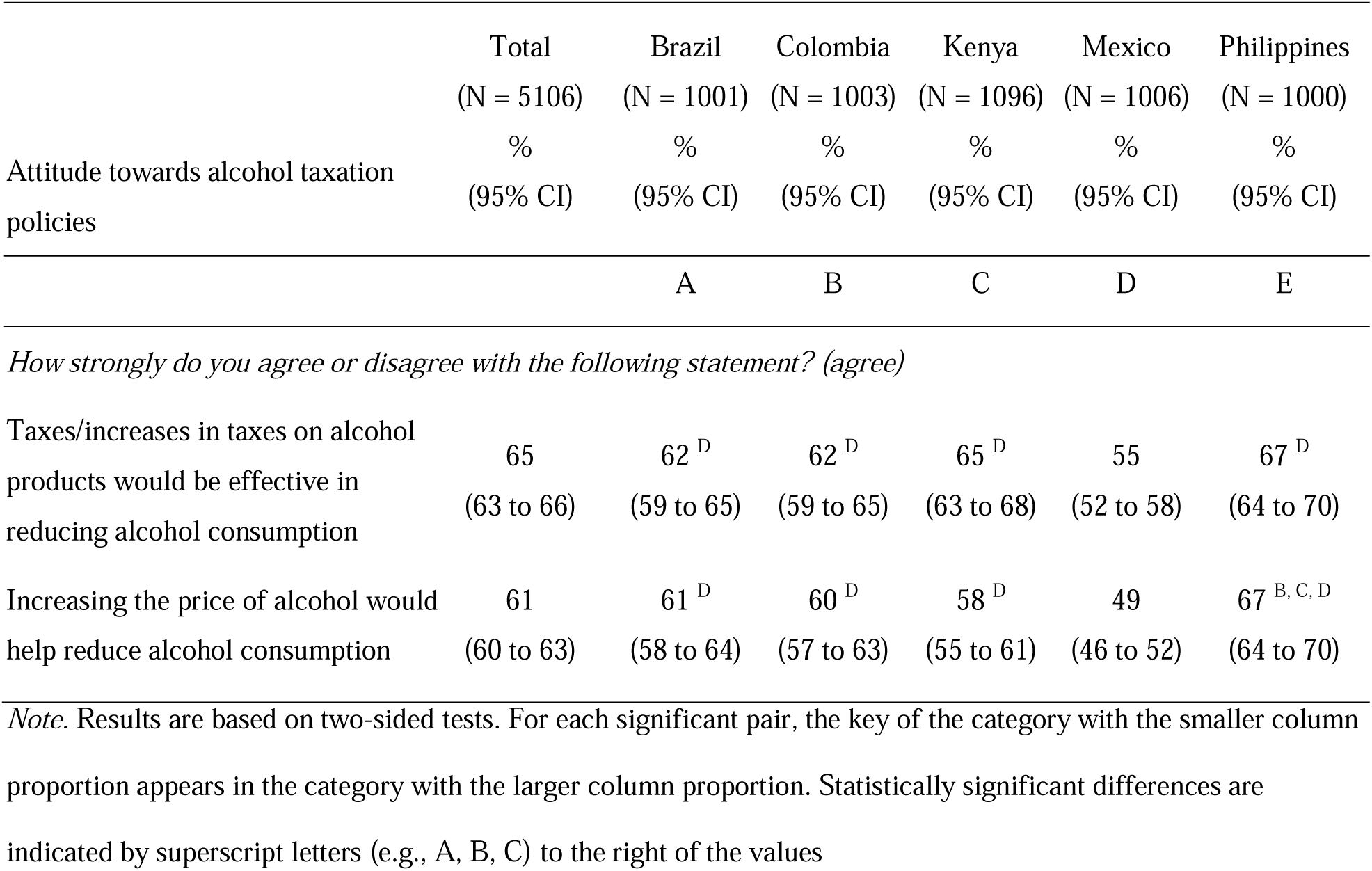
Attitude towards alcohol taxation policies by country.

### Predictors of agreement on the effectiveness of alcohol taxation policies

Table 5 presents the results of the ordinal regression analysis. Compared to participants in Brazil, those in Mexico and Kenya were significantly less likely, and those in the Philippines significantly more likely, to agree on the effectiveness of tax policy. Across countries, current alcohol consumers were less likely to agree than non-users. Gender, age and education level showed no statistically significant association.

**Table 5.** Predictors of perceived effectiveness of alcohol taxation policies (ordinal regression analysis)

|  | UOR | % 95 CI |  | p | AOR | % 95 CI |  | p |
| --- | --- | --- | --- | --- | --- | --- | --- | --- |
|  |  | LL | UL |  |  | LL | UL |  |
| *Country (ref: Brazil)* |  |  |  |  |  |  |  |  |
| Colombia | 0.91 | 0.77 | 1.07 | 0.25 | 0.90 | 0.73 | 1.11 | 0.33 |
| Kenya | 1.03 | 0.87 | 1.22 | 0.72 | 0.78 | 0.63 | 0.97 | 0.02* |
| Mexico | 0.65 | 0.56 | 0.78 | <0.01* | 0.68 | 0.55 | 0.83 | <0.01* |
| Philippines | 1.31 | 1.11 | 1.56 | <0.01* | 1.74 | 1.40 | 2.17 | <0.01* |
| *By what gender do you prefer to be known? (ref: Men)* |  |  |  |  |  |  |  |  |
| Women | 1.05 | 0.95 | 1.15 | 0.34 | 1.09 | 0.96 | 1.23 | 0.19 |
| *What is your age in completed years? (ref: 18 - 33 years)* |  |  |  |  |  |  |  |  |
| 34 - 49 years | 0.98 | 0.87 | 1.10 | 0.71 | 1.00 | 0.86 | 1.16 | 0.95 |
| 50 - 64 years | 0.91 | 0.8 | 1.03 | 0.13 | 0.86 | 0.73 | 1.02 | 0.09 |
| 65+ years | 0.94 | 0.79 | 1.11 | 0.45 | 1.13 | 0.89 | 1.45 | 0.31 |
| *What is the highest level of education you have completed? (ref: Primary)* |  |  |  |  |  |  |  |  |
| Secondary | 1.13 | 0.99 | 1.29 | 0.07 | 0.94 | 0.80 | 1.12 | 0.51 |
| Tertiary | 1.07 | 0.94 | 1.22 | 0.33 | 0.92 | 0.76 | 1.10 | 0.34 |
| *Have you ever consumed an alcohol such as beer, wine, spirits, gin, vodka or [add other local examples]? Alcohol user (ref: Never consumed alcohol)* |  |  |  |  |  |  |  |  |
| Former alcohol user | 0.80 | 0.71 | 0.89 | <0.01* | 0.95 | 0.81 | 1.13 | 0.57 |
|  |  | LL | UL |  |  | LL | UL |  |
| Current alcohol user | 0.59 | 0.52 | 0.66 | <0.01* | 0.74 | 0.62 | 0.89 | <0.01* |
| *In your opinion how much of a problem is alcohol in your country? (ref: not a problem/slight problem/moderate problem)* |  |  |  |  |  |  |  |  |
| Major problem | 1.32 | 1.20 | 1.46 | <0.01* | 1.16 | 1.01 | 1.33 | 0.03* |
| *During the last two months, have you seen, read or heard any advertisements, public service announcements or messages in the media about the harms related to alcohol consumption? (ref: No/don't know)* |  |  |  |  |  |  |  |  |
| Yes | 1.20 | 1.08 | 1.34 | <0.01* | 1.18 | 1.03 | 1.36 | 0.01* |
| *Knowledge about health or social harms of consuming alcohol (ref: low knowledge)* |  |  |  |  |  |  |  |  |
| Medium knowledge | 1.28 | 0.95 | 1.73 | 0.11 | 1.04 | 0.70 | 1.55 | 0.84 |
| High knowledge | 2.18 | 1.65 | 2.89 | <0.01* | 1.36 | 0.94 | 1.98 | 0.11 |
| *It is important to educate children and adolescents and youth about the risks and consequences of alcohol consumption (ref: disagree/neutral)* |  |  |  |  |  |  |  |  |
| Agree | 1.73 | 1.44 | 2.07 | <0.01* | 1.16 | 0.91 | 1.48 | 0.24 |
| *Alcohol easy to buy (ref: disagree/neutral)* |  |  |  |  |  |  |  |  |
| Agree | 1.22 | 1.07 | 1.39 | <0.01* | 1.04 | 0.88 | 1.22 | 0.66 |
| *Alcohol is relatively inexpensive in my country (ref: disagree/neutral)* |  |  |  |  |  |  |  |  |
| Agree | 1.50 | 1.35 | 1.67 | <0.01* | 1.4 | 1.24 | 1.59 | <0.01* |
| *It is the government's responsibility to address the problems with alcohol consumption in my country (ref: disagree/neutral)* |  |  |  |  |  |  |  |  |
| Agree | 2.51 | 2.22 | 2.83 | <0.01* | 1.70 | 1.47 | 1.97 | <0.01* |
| *Our current alcohol laws protect people from alcohol harms (ref: disagree/neutral)* |  |  |  |  |  |  |  |  |
| Agree | 0.7 | 0.64 | 0.77 | <0.01* | 0.81 | 0.82 | 0.92 | <0.01* |
| *Our current alcohol laws are poorly enforced/implemented (ref: disagree/neutral)* |  |  |  |  |  |  |  |  |
| Agree | 1.84 | 1.64 | 2.06 | <0.01* | 1.25 | 1.09 | 1.45 | <0.01* |
| *Policy measures to reduce the consumption of alcohol can benefit the public whether they consume alcohol or not (ref: disagree/neutral)* |  |  |  |  |  |  |  |  |
| Agree | 2.34 | 2.10 | 2.61 | <0.01* | 1.56 | 1.36 | 1.79 | <0.01* |
| *Alcohol companies can't be trusted to tell the truth about the harms of alcohol use (ref: disagree/neutral)* |  |  |  |  |  |  |  |  |
| Agree | 0.88 | 0.79 | 0.97 | 0.01* | 0.93 | 0.82 | 1.05 | 0.25 |
| *The alcohol industry interferes with the passage of new alcohol policies in my country (ref: disagree/neutral)* |  |  |  |  |  |  |  |  |
| Agree | 2.21 | 1.99 | 2.45 | <0.01* | 1.43 | 1.25 | 1.64 | <0.01* |
|  |  | LL | UL |  |  | LL | UL |  |
| *Alcohol companies should take responsibility for the harm caused by alcohol use (ref: disagree/neutral)<sup>2</sup>* |  |  |  |  |  |  |  |  |
| Agree | 2.04 | 1.83 | 2.28 | <0.01* | 1.46 | 1.28 | 1.66 | <0.01* |

*Note.* UOR, unadjusted odds ratio. AOR, adjusted odds ratio. Ref, reference category. LL, Lower level. UL, upper
level.
\* $p < .05$ .

Viewing alcohol as a major societal problem, perceiving it as inexpensive, and recent exposure to alcohol harm-related media messages were positively associated with agreement, whereas ease of alcohol purchase, general knowledge of alcohol harms, and views in favor of youth alcohol education were not significantly associated. Beliefs about government responsibility were positively associated with agreement on tax effectiveness, specifically: addressing alcohol harms is a government responsibility, policies benefit all, current laws protect people, and existing laws are poorly enforced. The belief that the alcohol industry interferes with policymaking and is culpable for alcohol-related damages was also positively associated with perceived tax effectiveness, but trust in alcohol companies showed no significant association.

## DISCUSSION

Our study reveals widespread public concern about alcohol as a societal issue across all countries, with violence-related harms consistently most prominent. Awareness of NCD-related harms was generally low. Among NCDs, liver cirrhosis was the most commonly cited concern, followed by kidney disease. Other NCDs—cancer, heart disease, stroke, and lung disease—were rarely mentioned. These findings highlight opportunities for policy advocacy to prioritize immediate public concerns about violence-related harms, while building greater awareness of chronic, long-term health risks to sustain support for alcohol policies (16).

Public concerns play a pivotal role in shaping support for alcohol policy. When alcohol is perceived as a broader societal threat—rather than a problem confined to a few individuals, like problem drinkers or pregnant women—there is stronger public support for population-level measures such as taxation, pricing and availability restrictions (16,22). Our finding that alcohol is widely viewed as a major societal problem provides an important foundation for advancing comprehensive alcohol policies. In particular, the prominence of violence-related harms in public consciousness suggests that framing alcohol policies around these highly salient concerns may serve as an effective entry point for mobilizing support and building sustained endorsement of population-level alcohol policies (22).

However, our findings also highlight a generally low level of public concern regarding alcohol’s contribution to non-communicable diseases (NCDs), including cancers, diabetes and stroke. This low level of awareness contrasts sharply with the substantial role these conditions play in the global burden of disease (1). This awareness gap may reflect a historical underemphasis on NCD-related harms in alcohol policy communication, compounded by persistent public uncertainty about causal links. From a public health perspective, this presents a critical challenge. Analogous experiences within public health, most notably in overdose prevention, demonstrate how framing these issues outside the health domain has often led to responses rooted in the enforcement or criminal justice systems rather than in prevention, care and community-based health strategies (16). When the health sector’s leadership is diminished or delayed, the effectiveness of policy responses may be compromised, and stigma can be reinforced, thereby weakening public engagement with lifesaving interventions. While intersectoral collaboration remains essential, neglecting the health dimension risks mischaracterizing the problem—and consequently, misdirecting the response (31). Our survey findings underscore the need for sustained public health communication to build health literacy about alcohol’s role in NCDs. Enhancing public understanding of these less visible but more pervasive harms is essential to securing long-term support for comprehensive, preventive alcohol policies.

Our study found that alcohol was generally perceived as relatively inexpensive and easily accessible across all countries. Public attitudes toward government action to curb consumption were also consistently favourable. Majorities in all countries affirmed that governments bear responsibility for addressing alcohol-related harms and expressed dissatisfaction with the adequacy and enforcement of current policies. In addition, participants in all countries, except Colombia, viewed the alcohol industry with scepticism, believing that companies should be held accountable for the harms associated with their products.

Across all countries, majorities agreed that alcohol taxation policies—such as removing financial incentives for alcohol companies and implementing alcohol taxes—are effective in reducing alcohol consumption. Previous research has demonstrated that believing a policy is effective corresponds to greater public support. (21,22) Our findings align with recent surveys measuring support for alcohol taxes. For example, while we found that 62% of Colombians agreed taxes are effective, a Gallup survey from 2021-22 reported 61% of Colombians supported higher alcohol taxes. (21,22,32) This consistency across surveys suggests clear public backing for alcohol taxation policies.

However, it is important to note that these supportive attitudes were expressed by relatively modest majorities, highlighting the fragility of public consensus and its vulnerability to disruption by vocal minority opposition. Evidence from behavioural science demonstrates that even when majority agreement appears strong, small but active minorities can still shift public opinion or stall policy progress, underscoring the need to build and reinforce public support (33). This was evident in related public health efforts to enact sugary beverage taxes in Mexico and the United States, where small but influential industry groups successfully disrupted or reversed policies despite public support (34–36).

Regression analyses identified key factors predicting stronger public support for alcohol taxes. The strongest predictors were believing that addressing alcohol harm is a government responsibility, recognizing that alcohol policies benefit everyone—not just drinkers—and believing that the alcohol industry interferes with or should be accountable for alcohol-related harms. Viewing alcohol as a major societal problem, perceiving alcohol as affordable, prior exposure to media messages on alcohol harms, and not currently drinking alcohol also predicted greater policy support. Demographic variables (gender, age, and education) and general knowledge of alcohol-related harms were not significant predictors.

These findings carry important implications for public health advocacy, particularly in how alcohol taxation is framed to build public support (22,37). That belief in government responsibility and industry culpability are strong predictors of perceived tax effectiveness suggests taxation is not seen merely as a neutral policy tool but as a mechanism for accountability (38) If taxes are viewed as punitive or coercive, support may weaken, especially among those who distrust government intervention or believe the industry can self-regulate. In such cases, industry-led solutions, like voluntary marketing codes or educational campaigns, may appear more appropriate. By contrast, when alcohol is framed as a societal problem beyond the reach of market mechanisms, and the industry is seen as a source of harm rather than a responsible actor, taxation becomes not only acceptable but necessary. It serves to internalize the external costs of alcohol-related harm, shifting the burden from the public to those who profit from its sale. This underscores the need to frame alcohol as a public issue requiring public solutions and to build confidence in the role of government and public health institutions. Equally important is exposing the limits of industry self-regulation. Together, these efforts are essential to expanding and sustaining public support for evidence-based alcohol taxation policies (39,40).

Finally, while there were broad universal patterns across countries, distinct country-level pictures of public perceptions and attitudes toward alcohol-related issues emerged from the study. Overall, the greatest concerns about alcohol harms and most favourable attitudes toward alcohol policies were observed in Kenya and Brazil, while the least were found in the Philippines. Respondents in Kenya showed the highest concern about alcohol harms and the strongest support for government responsibility and industry culpability. Respondents in Brazil highlighted health concerns, high alcohol accessibility and industry scepticism. In Mexico and Colombia, respondents emphasized violence-related harms but had lower health awareness, with people in Colombia notably positive toward the industry. Respondents in the Philippines had lowest overall concern yet strong support for alcohol taxation policies. This high support is likely influenced by the country’s prior experience with tobacco taxation and the visible use of sin tax revenues to expand universal health coverage—demonstrating how policy history can shape public expectations of similar or new fiscal measures. Importantly, this also suggests that public health communicators should actively draw from these historical and parallel experiences to build resonance and legitimacy for alcohol-related taxation policies. These country-level differences underscore the importance of tailoring advocacy and policy strategies to national contexts while leveraging shared regional patterns.

Our study has limitations common to opinion surveys, including self-report and social desirability biases. Non-response bias may affect generalizability, though we applied weighting and standardized measures to minimize this. The cross-sectional nature of the survey limits our ability to draw causal inferences. In addition, public opinion is dynamic, and the views captured may change over time. Nevertheless, our robust survey methods and adherence to best practices provide confidence in our findings, further validated by the consistency of our regression results with evidence from other studies.

## CONCLUSION

In conclusion, this study identifies shared and country-specific patterns in public attitudes toward alcohol harms and taxation. While alcohol is widely viewed as a societal problem, agreement on taxation effectiveness relies on beliefs about government responsibility and industry culpability. Empirical findings indicate immediate opportunities for policy action by addressing acute public concerns like violence, while highlighting gaps—particularly around chronic health risks—where sustained public mobilization is crucial. Framing alcohol as a public health and NCD prevention priority, referencing successful precedents like tobacco taxation, and reinforcing government responsibility will be key to advancing comprehensive alcohol policy reform.

## Acknowledgements

The authors gratefully acknowledge the contributions of the following partner organizations and colleagues who supported this study through leadership in project implementation, local coordination, and valuable inputs in finalizing the questionnaire:

- From Brazil: ACT Promoção da Saúde
- From Colombia: Fundación Anáas and Red PaPaz,
- From Kenya: International Institute for Legislative Affairs and Students’ Campaign Against Drugs (SCAD);
- From Mexico: El Poder del Consumidor, Salud Justa
- From the Philippines: Action for Economic Reforms
- Economic of Health at Johns Hopkins Bloomberg School of Public Health

We also thank the team at Vital Strategies — Bibiana Castillo, Tainá de Almeida Costa, Benjamin Gonzalez Rubio, Luyanda Majija, Patricia Baquiran, and Rebecca Perl — for their invaluable leadership and coordination throughout the research study.

We acknowledge Karen Schmidt for her editorial support in preparing the manuscript.

## Author contributions

NM: conceptualization and design of the work, supervision of data collection, design and direction of research materials, data analysis and interpretation, lead writer of all sections of this paper; NSN: contribution to design of this work, particularly research materials development, oversight of data collection, data curation, formal analysis, investigation, validation, writing, review and editing; RR: contribution to design of this work, particularly research materials development, oversight of data collection, data analysis, writing – review; MM: data modelling and analysis, validation, writing, review and editing; AJ: contribution to design of this work, particularly research materials development, data collection, data curation, formal analysis; JD: conceptualization of the work, data interpretation, review and editing of this manuscript; SM: conceptualization of the work, data interpretation, review and editing of this manuscript; AK: conceptualization and design of the work, supervision of data collection, design and direction of research materials, review and editing of this manuscript.

## Disclaimer

Open Philanthropy was not involved in any aspect of the writing of this manuscript. The authors have not received additional payment to write this article.

## Ethics approval

The study protocol and all materials were reviewed and approved by an Institutional Review Board through BRANY (Biomedical Research Alliance of New York), a U.S.-based IRB (Application Number: 2023-05-22-09-07). All participants received information outlining the purpose, procedures and voluntary nature of the study. Participation was entirely voluntary, and informed consent was implied by the completion of the survey. Participants were assured that their responses would remain anonymous and confidential, and they could withdraw from the study at any time without penalty.

## Data availability statement

Data are available upon request to first author (Nandita Murukutla; https://nmurukutla@vitalstrategies.org)

## Footnotes

1 Referred to within the narrative text as *culpability for alcohol-related damages* to more precisely characterize public perceptions of alcohol industry accountability.

2 Referred to within the narrative text as *culpability for alcohol-related damages* to more precisely characterize public perceptions of alcohol industry accountability.

